# Structural brain alterations and their associations with inattentive and hyperactive/impulsive behaviors show sex-differentiated patterns in young adults with chronic sports-related mild traumatic brain injury

**DOI:** 10.64898/2026.02.20.26346734

**Authors:** Ziyan Wu, Catherine A. Mazzola, Arlene Goodman, Yu Gao, Tara Alvarez, Xiaobo Li

## Abstract

Traumatic brain injury (TBI), particularly sports- and recreational activity related mild TBI (mTBI), is common in young adults and can be followed by persistent attentional and executive complaints. This study investigated chronic (≥6 months post-injury) structural brain alterations in gray matter (GM) and white matter (WM) and their associations with self-reported inattentive and hyperactive/impulsive symptoms, with a focus on sex-differentiated patterns. Structural brain properties in gray matter (GM) and white matter (WM) were acquired from 44 subjects with TBI and 45 matched controls, by utilizing structural MRI and diffusion tensor imaging techniques. Behavioral measures assessing severities of post TBI inattentive and hyperactive/impulsive symptoms were collected from each participant. Between-group and sex-specific differences of these brain and behavioral measures were conducted. Interactions among the TBI-induced significant brain- and behavioral-alterations, and their sex-specific patterns, were assessed as well. Male-dominated pattern of increased cortical thickness in superior parietal lobule (SPL) and female-dominated pattern of higher superior longitudinal fasciculus and superior fronto-occipital fasciculus (sFOF) fractional anisotropy (FA) were observed in the TBI group, when compared to controls. In males with TBI, greater SPL cortical thickness was significantly correlated with increased inattentive behaviors. In females with TBI, higher FA of sFOF was significantly correlated with decreased hyperactive/impulsive behaviors. Findings suggest that TBI-induced superior parietal cortical GM abnormalities may significantly cause attention deficits in patients with TBI, especially in males; while optimal post-TBI WM recovery in sFOF significantly contributes to maintenance of inhibitive control in patients with TBI, especially in females.

## Introduction

Traumatic Brain Injury (TBI) is one of the major public health concerns with approximately 70 million new cases occurring worldwide per year. ^1^ It is often caused by an external force to the head, such as hit, bump, or jolt, which can lead to both gray matter (GM) and white matter (WM) tissue damages. ^2^ Acute symptoms such as headache, dizziness, confusion, and fatigue following TBI may have quick and full recoveries. However, at least 15-30% of TBI subjects develop prolonged neurocognitive and behavioral impairments, such as attention and executive control deficits that significantly contribute to poor academic and social functioning and create lifetime challenges to the affected individuals. ^3–5^ Unfortunately, clinical studies found that such TBI-induced long-term neurocognitive and behavioral consequences have been treated based on symptom endorsements from subjective observations, and reported inconsistent results, with some showing effectiveness while many others not. ^6–8^ It is critical to understand TBI-associated alterations in brain and behavior and their interactions, so that more refined long-term treatment and intervention strategies can be developed.

Over the last two decades, increasing neuroimaging studies have reported structural and functional brain alterations and their associations with neurocognitive and behavioral impairments in individuals with TBI. A voxel-based morphometry study reported significantly reduced GM concentration in frontal, temporal and parietal cortices and their significant associations with attention deficits in young adults with TBI. ^9^ Significant GM volumetric atrophy in cingulate gyrus was reported in adults with TBI, without showing linkages with neurocognitive and behavioral impairments in the TBI subjects. ^10^ An early diffusion tensor imaging (DTI) study linked increased fractional anisotropy (FA) of the anterior corona radiata to better ability for attention switch in adults with mild TBI (mTBI). ^11^ Significantly higher FA of the posterior corpus callosum was observed in patients with mTBI relative to matched controls, and was linked with poorer inhibitory control capacity. ^12^ Besides these, structural alterations in other brain regions, such as temporal lobe, thalamus, hippocampus and fornix, were reported and found to link with cognitive/behavioral impairments related to decision making, processing speed, visual tracking and working memory, in adolescents and adults with TBI. ^13–16^ However, a recent DTI study reported no significant WM abnormality in athletes after sports-related concussion, relative to matched controls. ^17^ Meanwhile, an early functional MRI study reported significantly reduced neural activations in posterior parietal cortex, frontal eye fields and ventrolateral prefrontal cortex during attentional disengagement in adults with mTBI, relative to group-matched controls. ^18^ Increased resting-state functional connectivity in the posterior cingulate gyrus and decreased connectivity in frontal and parietal regions were reported in adolescent athletes with concussion, relative to controls. ^19^ Our recent functional near-infrared spectroscopy study reported significantly increased functional connectivity between right inferior occipital cortex and bilateral calcarine gyri in adults with TBI during a visual sustained attention task, which were significantly associated with inattentive and hyperactive/impulsive behaviors in TBI subjects relative to matched controls. ^20^ Greater activation in the posterior cerebellum and its association with additional demand for inhibitory control was reported in a functional MRI study in children with mTBI, when compared to matched controls. ^21^ Furthermore, altered hemodynamic responses and functional connectivities associated with temporal, insular cortices, and subcortical areas were also demonstrated in children and adults with TBI relative to controls in other functional MRI studies. ^22–24^ The significant inconsistency of findings from the existing structural and functional neuroimaging studies in TBI can be partially resulted from technical differences in data acquisitions and analyses, and sample-related biases such as differences in injury mechanism, post-injury stage, varying age ranges, and sex ratios.

Indeed, sex-specific patterns of TBI-induced cognitive and behavioral impairments have been largely documented, ^25–34^ resulting in contradictory results with some reported significantly worse performances of visual, verbal memory and executive control domains in females with TBI, relative to male TBI patients, ^25–30^ while others demonstrated better performances in females ^31–34^ or no substantial sex differences. ^35^ Several existing neuroimaging studies have attempted to investigate sex-related brain mechanisms associated with TBI. ^36–41^ Specifically, a DTI study reported significantly decreased FA in bilateral uncinate fasciculi in males with TBI relative to females TBI patients and controls. ^36^ A structural MRI study found significantly increased cortical thickness in the left caudal anterior cingulate cortex in females with TBI, compared to male patients. ^39^ A resting-state functional MRI study reported significantly increased connectivity between the orbitofrontal cortex (OFC) and parietal, occipital and cerebellum areas as well as significantly decreased connectivity between the OFC and temporal, insular regions in females with TBI relative to males TBI patients. ^38^ Another resting-state functional MRI study found increased intrinsic functional connectivity in motor network, ventral stream network, executive function network, cerebellum network and decreased connectivity in visual network in males with TBI relative to females TBI patients. ^41^ Again, findings from these investigations on sex-related brain alterations in TBI are far from converging.

Motivated to further investigate the interactions between TBI-induced structural brain alterations and cognitive/behavioral impairments, as well as their sex-specific patterns, structural MRI and DTI data from 44 young adults with TBI and 45 matched normal controls (NC) were acquired for this study. Behavioral measures including inattention and hyperactivity/impulsivity, which were among the most frequently reported domains impacted by TBI, ^42–44^ were also collected from every subject. Sex distributions were balanced in both groups. To further minimize sample-related bias, all the TBI subjects were recruited from sports teams in New Jersey Institute of Technology (NJIT) and Rutgers University (Rutgers), with clinically confirmed sports- and/or recreational activity-related TBIs at least 6 months before the study date, and aged 18-27 years old. Based on findings of previous studies conducted by our team and other researchers, ^9, 18, 20, 38^ we hypothesized that injury induced structural alterations associated with frontal, parietal and occipital cortices, regions well-recognized to play important roles in attentional and inhibitory deployment, would significantly contribute to inattentive and hyperactive/impulsive behaviors in affected individuals. We further expected that these structural brain abnormalities would demonstrate sex-specific patterns.

## Materials and Methods

### Participants

A total of 89 young adults (ranging from 18 through 27 years of age) were initially involved in this study. Forty-four were TBI patients, including 23 male TBI patients and 21 female TBI patients, and 45 were group-matched NC, including 23 males and 22 females. Participants of both groups were recruited from NJIT and Rutgers, with patients from sports teams and controls through study flyers. The study received Institutional Reviewed Board Approvals at NJIT and Rutgers. Written informed consents were provided by all participants. Demographic characteristics were summarized in **Table 1**.

**Table 1:** Demographic and clinical characteristics in the groups of NC and TBI.

|  | NC<br>(N=45) | TBI<br>(N=44) |  |
| --- | --- | --- | --- |
|  | Mean (SD) | Mean (SD) | *p* |
| Age | 22.25 (2.72) | 21.58 (1.97) | .186 |
| Education year | 14.91 (1.95) | 14.20 (1.55) | .064 |
| Mother's education year | 15.20 (2.63) | 15.64 (2.72) | .450 |
| Father's education year | 15.53 (3.18) | 15.53 (2.75) | 1.00 |
| CAARS scores |  |  |  |
| Inattentive raw scores | 4.64 (2.76) | 9.25 (6.14) | <.001 |
| Inattentive T-scores | 45.76 (6.38) | 56.98 (14.84) | <.001 |
| Hyperactive/impulsive raw scores | 5.07 (2.71) | 9.27 (5.68) | <.001 |
| Hyperactive/impulsive T-scores | 42.51 (5.81) | 52.80 (14.38) | <.001 |
|  | N (%) | N (%) | *p* |
| Male | 23 (51.11) | 23 (52.27) | .913 |
| Right-handed | 45 (100) | 44 (100) | 1.00 |
| Race/ Ethnicity |  |  | .219 |
| Caucasian | 14 (31.91) | 21 (46.67) |  |
| Black or African American | 4 (8.51) | 7 (17.78) |  |
| Asian | 20 (44.68) | 10 (22.22) |  |
| Hispanic/Latino | 3 (6.38) | 2 (4.44) |  |
| More than one race | 4 (8.51) | 4 (8.88) |  |

NC: normal controls; TBI: traumatic brain injury; N: number of subjects; SD: standard deviation; *p*: level of significance; CAARS: Conner's Adult ADHD Self-Reporting Rating Scales.

Specific inclusion criteria for the patient group were: having a history of one or multiple sports- and/or recreational activity-related TBIs clinically confirmed at least 6 months prior to study enrollment; having no head injury which caused overt focal brain damage; having no history of diagnosis with attention-deficit/hyperactivity disorder (ADHD) prior to the first onset TBI. Specific inclusion criteria for NC were: having no history of TBI; having no history of diagnosed ADHD; T-score < 60 for both the inattentiveness and hyperactivity/impulsivity subscales in Conner’s Adult ADHD Self-Reporting Rating Scales (CAARS), ^45^ which were administered during study assessments. General inclusion criteria for both subject groups were: native or fluent speakers of English, strongly right-handed measured using Edinburgh Handedness Inventory. ^46^ General exclusion criteria for both groups were: having a history or current diagnosis of neurological and psychiatric disorders; received treatment with any non-stimulant psychotropic medication within the month prior to testing; MRI constraints such as metal implants, claustrophobia, etc.

During the study, the general demographical information, current status and history of medical conditions were gathered from each participant. The Edinburgh Handedness Inventory and CAARS were administered. Because post-TBI attentional difficulties often extend beyond inattention to include inhibitory control complaints, such as impulsivity and difficulty suppressing prepotent responses, which are commonly linked to fronto-parietal control circuitry, we also evaluated hyperactive/impulsive symptoms. Leveraging CAARS subscales allowed us to quantify these two complementary attention-related dimensions within a single, well-validated instrument, providing a more complete characterization of attention and self-regulatory symptoms relevant to TBI. Moreover, structural MRI and DTI data were acquired. In our study, one subject in the TBI group had been taking short-acting psychostimulants for treatment of inattentive symptoms, and was instructed to have a wash-out period for at least 24-hour prior to the MRI scanning procedure.

After individual-level neuroimaging data analyses, 4 subjects were excluded from further data analyses due to excessive head motions (with any of the 6 realignment parameters > 2.5 mm). Therefore, a total of 85 subjects (including 43 controls and 42 TBI patients) were involved in group-level analyses.

### MRI data acquisition protocol

The T1-weighted structural MRI and DTI scans from each subject were acquired from a 3.0T Siemens Trio imaging system (Siemens, Erlangen, Germany). The structural MRI data were collected using a magnetization prepared rapid gradient echo sequence with the following parameters: repetition time (TR) = 1900 ms, echo time (TE) = 2.52 ms, flip angle = 9°; field of view (FOV) = 350mm×263mm×350 mm, voxel size = 1.0mm×1.0mm ×1.0mm, number of slices = 176. DTI data were acquired using an echo planar imaging pulse sequence with the following parameters: TR= 7700 ms, TE= 103 ms, FOV = 220mm×220mm×138 mm, voxel size = 2.0mm×2.0mm ×2.5mm, number of slices = 55, b-value = 700 s/mm^2^ along 30 independent directions, as well as a reference volume without diffusion-weighting (b = 0 s/mm^2^).

### Individual-level neuroimaging data analyses

For each subject, structural MRI data was motion corrected and then processed using an automated surface reconstruction model in the FreeSurfer v.6.0 software package (https://surfer.nmr.mgh.harvard.edu/). All of the steps were done using the recommended standard parameters. ^47^ To estimate regional cortical thickness and surface area, non-brain tissue was removed using a hybrid watershed/surface deformation procedure. Each volume was registered to the Talairach atlas ^48^ using an affine registration method. Intensity variations caused by magnetic field inhomogeneities were corrected. A cutting plane was defined to separate the left and right hemispheres and to remove the cerebellum and brain stem. Two surfaces between the GM and WM (called WM surface) and between the GM and cerebrospinal fluid (called pial surface) were generated using the triangular tessellation technique. The GM/WM border surface was then inflated to an average spherical surface to locate both the pial surface and the GM/WM boundary. Cortical thickness was measured as the average of two distances including the distance from each white surface vertex to their corresponding closet point on the pial surface and vice versa. Surface area was quantified by averaging the surrounding triangular face of the surface representation with vertex coordinates. Cortical parcellation was provided based on the Desikan-Killiany atlas. ^49^ According to our hypotheses, cortical GM regions of interest (ROIs) included all of the subregions of bilateral frontal (22 subregions), parietal (10 subregions), and occipital (8 subregions) cortices. Cortical thickness and surface area of these 40 bilateral cortical ROIs were included in group-level analyses.

DTI data of each subject was first preprocessed using the Diffusion Toolbox from FMRIB Software Library (FSL) version 5.0.9. ^50^ Eddy-current induced geometrical distortions and head motion were estimated and corrected. Voxel-based whole-brain FA maps were then created by fitting a diffusion tensor model. For DTI data, ROI-based and tractography-based analyses were conducted in each subject.

In the ROI-based analysis of each DTI data, each voxel-based FA map was first non-linearly aligned to the 1×1×1 mm MNI152 space (a normalized and averaged brain atlas developed by the Montreal Neurologic Institute). According to our hypotheses, four WM ROIs were extracted, including bilateral superior longitudinal fasciculus (SLF) (which connects frontal, parietal and occipital lobes), and superior fronto-occipital fasciculus (sFOF) (which interconnects frontal and occipital lobes), based on the Johns Hopkins University human brain WM tractography atlas, ^51^ and then mapped to each individual’s voxel-based FA image. **Figure 1A** demonstrated the SLF and sFOF ROIs in the left hemisphere. The average FA of each WM ROI was included in group-level analyses.

**Figure 1:**
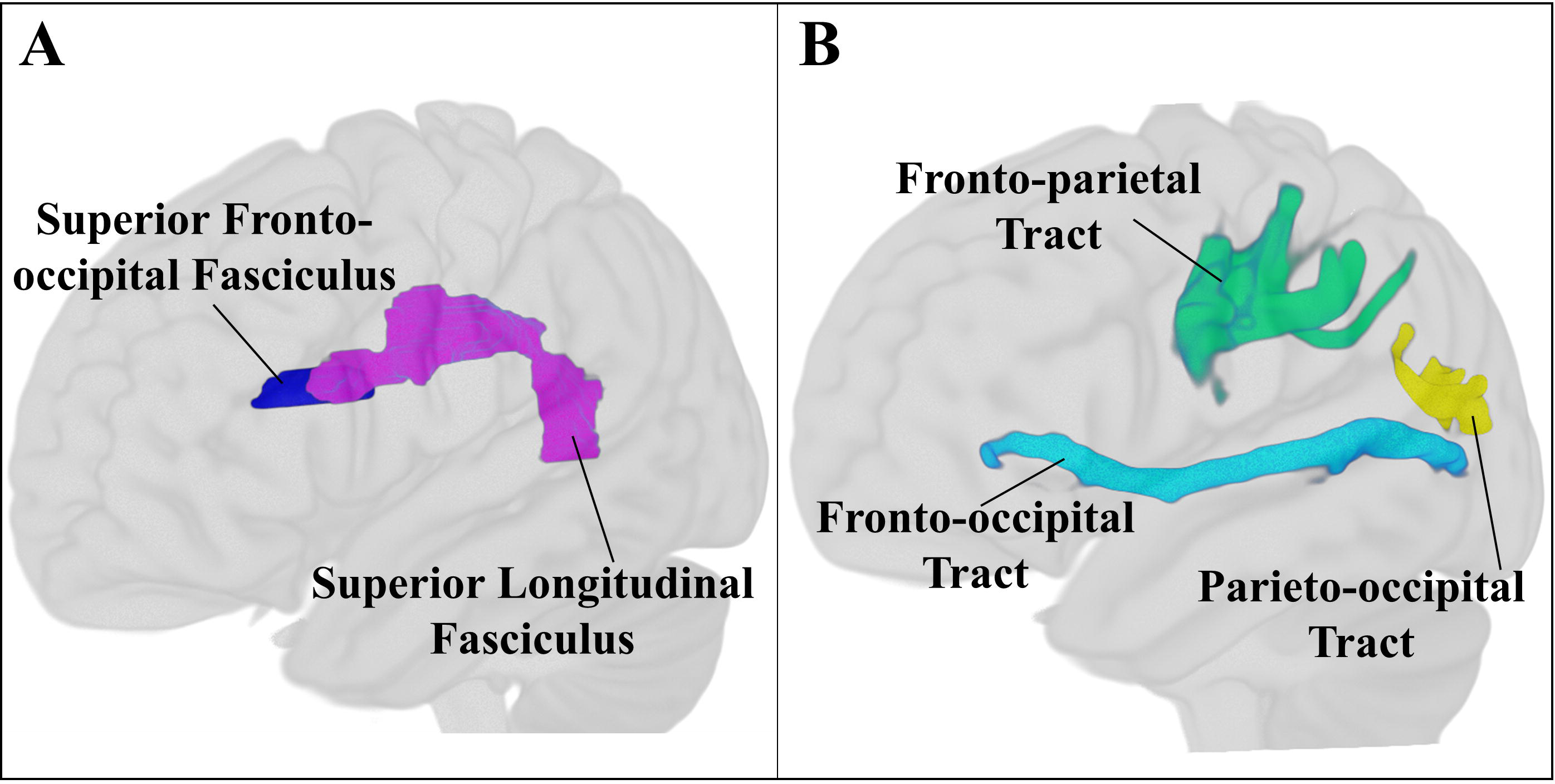
(A) White matter (WM) regions-of-interest selected based on Johns Hopkins University human brain WM tractography atlas; (B) WM tracts generated from probabilistic tractography analysis (displayed only in the left hemisphere).

In the tractography-based analysis of each DTI data, three cortical seeds were first determined in each hemisphere, including the frontal, parietal and occipital cortices parcellated from the structural MRI data of this subject using Desikan-Killiany atlas, ^49^ and then linearly registered to the native diffusion space. Within each voxel of each seed, two crossing fibers were estimated using the FSL/BEDPOSTX toolbox. ^52^ Probabilistic tractography in each pair of the three seeds was conducted with the following parameters: 5000 individual pathways were drawn on the principle fiber direction of each seed voxel within the ROI, with a step length of 0.5mm and maximum travel steps of 2000 for each sample pathway. Curvature threshold of 0.2 was set to exclude implausible pathways. **Figure 1B** depicted the three WM tracts in left hemisphere generated in this step. The FA and volume of each tract were extracted and involved in the group-level analyses.

### Group statistical analyses

The demographical and clinical variables were compared between controls and TBI patients, and then between males and females in each diagnostic group, using chi-square tests for discrete variables (i.e. sex) and unpaired two-sample t-tests for continuous variables.

Neuroimaging measures (including cortical thickness and surface area of 40 bilateral cortical GM ROIs, FA of the 4 WM ROIs and 6 tractography-based WM tracts) were compared using one-way analysis of covariance between controls and TBI patients, by controlling age, participant’s and their parents’ education levels as covariates. For the anatomical measures which showed significantly between-group differences, post-hoc t-tests were further conducted to examine sex-specific comparisons between the groups of TBI and controls. For each analysis, the threshold of significant difference, *p* ≤ 0.05, was determined after controlling multiple comparisons using the false discovery rate. ^53^

Furthermore, partial correlation analyses were conducted between the inattentive and hyperactive/impulsive symptom scores and the neuroimaging measures which showed significant between-group and sex-specific differences, by controlling participant’s age, education level and their parents’ education levels as covariates.

## Results

### Demographic and clinical characteristics

Relative to NC, TBI patients showed significantly higher raw and T scores for inattentive and hyperactive/impulsive symptoms measured by the CAARS subscales, while no between-group differences were observed in any demographic measures (**Table 1**). In addition, between-sex comparisons in the NC and TBI groups did not show significant differences in any clinical/demographic measures.

### Neuroimaging measures

As shown in **Tables 2** and **3**, relative to controls, TBI patients had significantly increased regional cortical thickness in the right superior parietal lobule (SPL) (F=6.954, *p*=0.050). Post-hoc analyses further showed that males with TBI had significantly increased cortical thickness of right SPL when compared to male controls (t=2.729, *p*=0.009).

**Table 2:** Neuroanatomical measures which showed significant differences between the groups of NC and TBI.

|  | Neuroanatomical Measures | NC<br>Mean (SD) | TBI<br>Mean (SD) | F-value | *p*-value after<br>FDR correction |
| --- | --- | --- | --- | --- | --- |
| NC<br>v.s.<br>TBI | CT of right superior parietal lobule | 2.278 (0.121) | 2.347 (0.131) | 6.954 | **0.050** |
|  | FA of left superior longitudinal fasciculus ROI | 0.504 (0.035) | 0.518 (0.020) | 5.137 | **0.039** |
|  | FA of left superior fronto-occipital fasciculus ROI | 0.539 (0.044) | 0.565 (0.042) | 5.820 | **0.039** |

NC: normal controls; TBI: traumatic brain injury; SD: standard deviation; CT: regional cortical thickness; FA: fractional anisotropy; FDR: false discovery rate.

**Table 3:** Neuroanatomical measures which showed significant or a trend of significant between-group differences in males and females.

|  | Neuroanatomical Measures | NC<br>Mean (SD) | TBI<br>Mean (SD) | t-value | p-value after<br>FDR correction |
| --- | --- | --- | --- | --- | --- |
| NC_M<br>v.s.<br>TBI_M | CT of right superior parietal lobule | 2.273 (0.149) | 2.385 (0.130) | -2.729 | **0.009** |
|  | FA of left superior longitudinal fasciculus ROI | 0.514 (0.034) | 0.528 (0.017) | -1.688 | 0.101 |
|  | FA of left superior fronto-occipital fasciculus ROI | 0.545 (0.047) | 0.571 (0.041) | -1.959 | 0.101 |
| NC_F<br>v.s.<br>TBI_F | CT of right superior parietal lobule | 2.283 (0.086) | 2.304 (0.121) | -0.679 | 0.501 |
|  | FA of left superior longitudinal fasciculus ROI | 0.491 (0.034) | 0.509 (0.017) | -2.048 | **0.053<sup>†</sup>** |
|  | FA of left superior fronto-occipital fasciculus ROI | 0.532 (0.041) | 0.559 (0.043) | -1.997 | **0.053<sup>†</sup>** |

NC\_M: male normal controls; TBI\_M: male traumatic brain injury patients; NC\_F: female normal controls ; TBI\_F: female traumatic brain injury patients; SD: standard deviation; CT: regional cortical thickness; FA: fractional anisotropy; FDR: false discovery rate; <sup>†</sup>: trend of significant difference.

Meanwhile, TBI patients showed significantly higher FA of the left SLF ROI (F=5.137, *p*=0.039), when compared to controls. Post-hoc analyses further showed that females with TBI had a trend of significantly higher FA of the left SLF ROI (t=2.048, *p*=0.053) compared to females in the NC group. Compared to controls, the TBI group also showed significantly higher FA of the left sFOF (F=5.820, *p*=0.039). We did not observe between-group differences in the 6 tractography-based WM measures.

### Brain-behavioral associations

As shown in **Figure 2**, greater regional cortical thickness of right SPL was significantly correlated with increased inattentiveness measured by the CAARS inattentive subscale in all male participants (r=0.389, *p*=0.011), especially in males of the TBI group (r=0.629, *p*=0.004). Meanwhile, higher FA of the left sFOF ROI was significantly correlated with decreased hyperactive/impulsive behaviors measured by the CAARS hyperactive/impulsive subscale in females of the patient group (r=-0.629, *p*=0.007).

**Figure 2:**
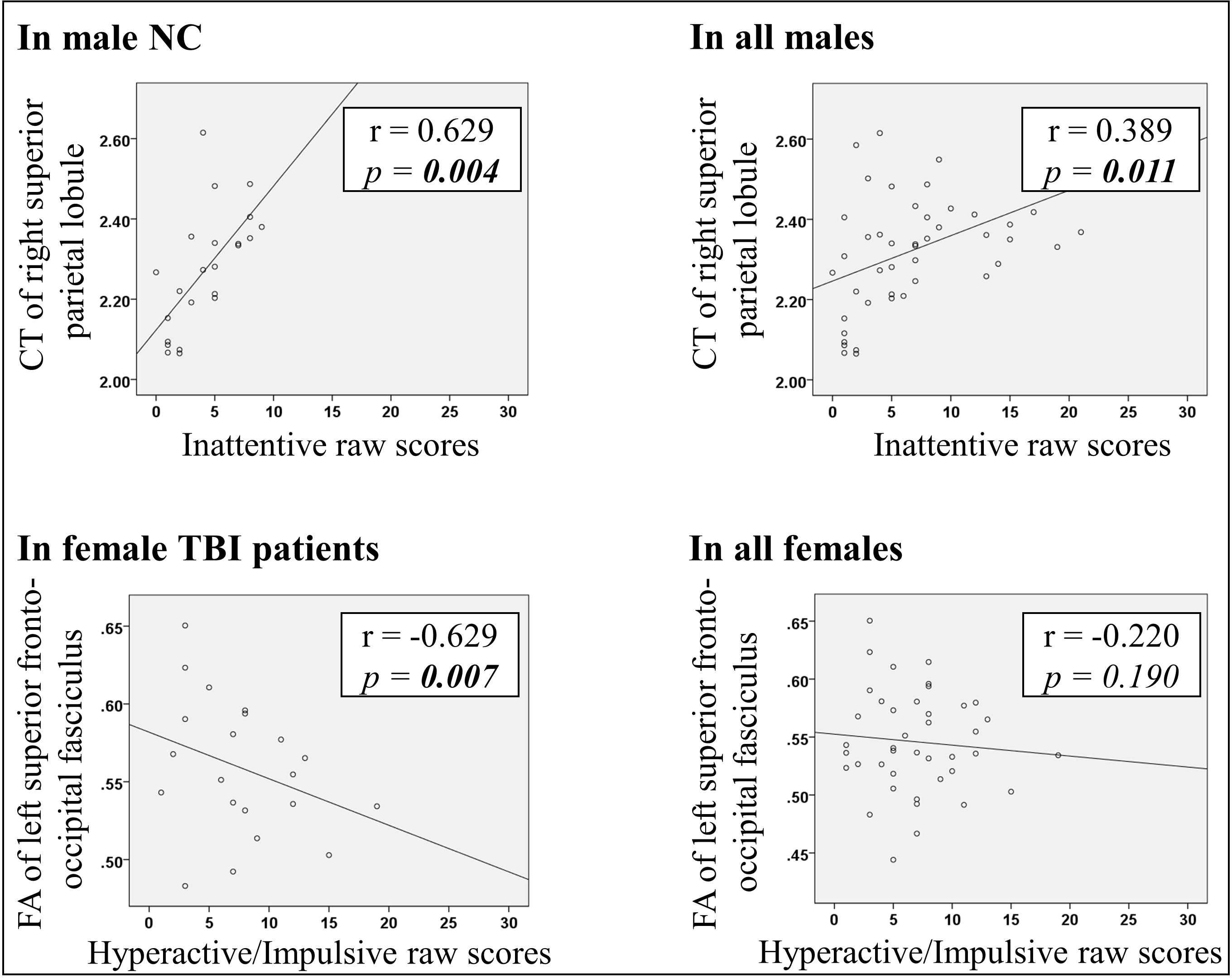
Associations between neuroanatomical measures and inattentive and hyperactive/impulsive symptom scores. (NC: normal controls; TBI: traumatic brain injury; CT: cortical thickness; FA: fractional anisotropy; r: strength of correlation; *p*: level of significance)

## Discussion

This study examined structural correlates of attention-related symptom dimensions in a restricted cohort of young adults with chronic sports/recreational mTBI and matched controls, using complementary GM (cortical thickness/surface area) and WM (FA and tractography) measures. By using a homogeneous injury mechanism, chronic stage definition, and balanced sex distribution, the findings provide evidence for sex-differentiated structural patterns and sex-specific symptom associations within attention-related domains.

We observed significantly increased cortical thickness in right SPL in the TBI group, when compared to controls. Structural alterations in parietal regions have been consistently reported in previous TBI studies. Recent structural MRI studies reported significantly increased cortical thickness of medial SPL in patients with chronic mTBI, ^39, 54^ consistently higher parietal cortical thickness in patients with acute-through-chronic mTBI (over a 3-month interval since the onset of brain injury), when compared to group-matched controls. ^55^ Although the biological mechanisms of such phenomena are still unclear, the underlying cellular process during the post TBI neuroinflammation stage may partially contribute to increased cortical thickness in affected brain regions. ^56, 57^

Besides the main finding of significantly increased cortical thickness in right SPL in the group of TBI, our post hoc analyses found that this TBI-higher-than-NC pattern regarding to cortical thickness of right SPL was mainly driven by the extremely greater right SPL cortical thickness in males with TBI relative to males in the NC group. Meanwhile, we found significant correlation between greater regional cortical thickness of right SPL and increased inattentiveness in males of the entire sample, but not in females. SPL is a major subregion of posterior parietal cortex, which plays an important role in sensory information transformation and attentional modulation. ^58–62^ Structural alterations associated with this region have been found to contribute to visual attentional disorders such as neglect. ^58, 63^ Functional neuroimaging studies in patients with TBI have reported increased SPL activation during attention processing ^64, 65^ and increased functional connectivity within the parietal lobe during resting state. ^66^ Although sex differences associated with SPL structural alterations have not been reported in human-based TBI studies, animal studies have consistently observed more aggressive neuroinflammatory profile in male mice compared to female mice in multiple brain injury models. ^67, 68^ Together with findings from these existing studies, our results may further suggest that compared to that in females, TBI-induced cortical abnormalities in SPL are more vulnerable to contribute to increased inattentive behaviors in males.

Results of the current study also showed that compared to the controls, subjects with TBI had significantly higher FA of left SLF and sFOF. SLF is a large bundle of association fibers in each hemisphere connecting the parietal, occipital and temporal lobes with ipsilateral frontal cortices, which is an essential component for higher level cognitive processes; ^69^ while sFOF is initiated from a compact fascicle at the level of the anterior horn of the lateral ventricle and terminated at the parietal region via the lower part of caudate, ^70^ and was suggested to play an important role in inhibitory control. ^71^ Increased FA in SLF was previously reported in youth with TBI, when compared to controls. ^72^ Although its biological mechanism is still unclear, studies suggested that prolonged TBI-reactive WM intracellular processes may partially stimulate axon regenerations, and result in increased FA. ^73–75^

In addition, the sex-specific post hoc analyses further found that the TBI-higher-than-NC pattern regarding to FA of left sFOF and left SLF, respectively, were both mainly led by higher FA of the ROIs in females with TBI relative to females in the NC group. And higher FA of left sFOF was significantly correlated with decreased hyperactive/impulsive behaviors in females of the patient group. So far, only a few studies investigated sex-differences in TBI-induced structural brain alterations and their associations with the clinical/behavioral outcomes, related to ability of processing speed, working memory and executive function. ^36, 39, 40^ Although there is lack of directly comparable results, previous studies have consistently observed strong associations between increased regional WM FA and better performance on processing speed, and awareness in TBI patients. ^76–78^ Adding into existing literature, our findings suggest that improved WM integrity in left SLF and sFOF can happen due to TBI-reactive WM intracellular processes and axon regenerations, which may partially modulate the inhibitory control function and contribute to better inhibitive behaviors, especially in females with TBI.

There are some issues of this study that need to be further discussed. First, this study assessed both group-wise and sex-specific patterns of the TBI-related structural brain alterations. Although a total of 85 subjects (with both sex included) involved in group-level comparisons, each post hoc sex-specific analysis was conducted within roughly a half of the entire sample, which was a relatively smaller sample size and may cause reduced statistical power. Future studies can focus on investigating sex effect in TBI, by recruiting a larger study sample. Second, one participant with TBI had been taking short-acting medications for inattentive symptoms, while all others were medication naive. An at least 24-hour wash-out period was instructed to this subject, before the MRI procedure. Nevertheless, there is no evidence from existing studies demonstrating significant impact of any short-acting stimulants to GM or WM brain structures. ^79^

In summary, this study demonstrated male-dominated pattern of significantly increased right SPL cortical thickness and female-dominated pattern of significantly higher FA in the left SLF and sFOF in patients with TBI, when compared to controls. Moreover, TBI-induced cortical abnormalities in SPL may significantly contribute to increased inattentive behaviors in males, while increased WM integrity in left SLF and sFOF may play an important role in better inhibitive control in females with TBI.

## Data Availability

The data that support the findings of this study are available from the corresponding author upon reasonable request. Due to ethical and privacy restrictions involving human participants, the data are not publicly available.

## Acknowledgments

This work was partially supported by research grants from the New Jersey Commission on Brain Injury Research (CBIR17PIL012), the National Institutes of Mental Health (R03MH109791, R15MH117368, and R01MH060698), the New Jersey Institute of Technology Start-up Award to Dr. Xiaobo Li.

## Author Disclosure Statement

All authors claim that no competing financial interests exist.

## Notes

### Competing Interest Statement

The authors have declared no competing interest.

### Author Declarations

The Institutional Review Board of New Jersey Institute of Technology and the Institutional Review Board of Rutgers, gave ethical approval for this work. Written informed consent was obtained from all participants.

